# Safety and efficacy of management for intraprocedural rupture during endovascular treatment for intracranial aneurysms

**DOI:** 10.1101/2024.01.23.24301699

**Authors:** Sakyo Hirai, Ryoichi Hanazawa, Masataka Yoshimura, Keigo Shigeta, Yohei Sato, Naoki Taira, Yoshihisa Kawano, Jun Karakama, Yoshiki Obata, Mutsuya Hara, Kenji Yamada, Yosuke Ishii, Kana Sawada, Shogo Imae, Hikaru Wakabayashi, Hirotaka Sagawa, Kyohei Fujita, Shoko Fujii, Satoru Takahashi, Akihiro Hirakawa, Shigeru Nemoto, Kazutaka Sumita

## Abstract

**Background:** Although intraprocedural rupture (IPR) is rare, it is a devastating complication of endovascular treatment (EVT) for intracranial aneurysms. Very few studies have been conducted on IPR, and the safety and efficacy of management techniques of IPR have not been investigated.

**Methods:** Patients who underwent IPR during EVT between 2013 and 2022 were enrolled from the multicenter observational registry. Focusing on the management of IPR, we examined its safety and efficacy using imaging markers, including increased hemorrhage and ischemic lesions evaluated using postoperative computed tomography and diffusion-weighted imaging, respectively.

**Results:** Of the 3269 EVT for intracranial aneurysms, 74 patients who underwent IPR (2.26%) were analyzed. Fifty-five patients (3.36%) experienced IPR in 1636 EVT cases for ruptured aneurysms. The multivariate analysis revealed that increased hemorrhage was significantly associated with poor outcomes (odds ratio [OR], 6.67 [95% confidence interval (CI), 1.07–41.44], p=0.042), whereas ischemic lesions were not. Regarding management techniques of IPR, antihypertensive medication use was significantly associated with increased hemorrhage (OR, 13.17 [95% CI, 2.26–76.69], p=0.004). Heparin reversal was an independent factor for ischemic lesions (OR, 4.81 [95% CI, 1.09–21.14], p=0.038).

**Conclusions:** Even though the setting of IPR may be miscellaneous, and optimal management varies depending on individual cases, heparin reversal might be associated with ischemic complications rather than being useful for controlling bleeding in IPR during EVT for ruptured aneurysms.

## Introduction

Although recent technological advances in endovascular treatment (EVT) for intracranial aneurysms have yielded improved clinical outcomes, intraprocedural rupture (IPR) remains one of the most devastating complications. The incidence of IPR during EVT is 2–6%; however, the IPR-related morbidity and mortality rates account for up to 60% ^1–4^. However, some reports suggest that IPR of unruptured aneurysms may lead to significantly favorable outcomes with appropriate management ^5, 6^. As IPR can be an uncontrollable event with a certain probability, its appropriate management is crucial in practical clinical settings ^7–9^.

Various management strategies for IPR, including heparin reversal, antihypertensive drug administration, and endovascular procedures using balloon catheters, have been reported ^3, 10, 11^. Although these conventional methods are theoretically presumed to be effective in daily clinical practice, they have not yet been scientifically verified. Given the priority placed on hemostasis, there may be a lack of consideration of the adverse effects of IPR management, such as thromboembolic complications associated with balloon catheter inflation or heparin reversal. Moreover, very few studies have verified the effect of these management techniques for IPR on clinical outcomes because of its infrequency, and its urgency makes it extremely difficult to examine them in prospective studies.

The primary purpose of the present study was to review our experience with IPR during EVT for intracranial aneurysms and to clarify its clinical characteristics. Second, we investigated the safety and efficacy of conventional management strategies for IPR.

## Methods

### Patient Selection

This retrospective, multicenter, observational cohort study was conducted at 14 stroke centers in Japan using data from the EVT registry. The registry retrospectively (before 2021) and prospectively (after 2021) enrolled patients who underwent elective or emergent EVTs at the participating institutions. The ethics committees of the participating institutions approved this registry (Approved number: M2020-102). Written informed consent was waived because the study was retrospective with no additional invasive procedures or costs for the participants, and the information was sufficiently anonymized. The data supporting the findings of this study are available from the corresponding author upon reasonable request. This study was conducted in accordance with the Strengthening the Reporting of Observational Studies in Epidemiology (STROBE) Statement. Patients who underwent EVT for intracranial aneurysms and experienced IPR between January 2013 and December 2022 were included using data from the EVT registry.

### Clinical Data Collection

Before collecting clinical data retrospectively, a uniform definition of each clinical measurement and the protocols of imaging analyses were agreed upon via meetings attended by researchers at the participating stroke centers.

The following data were collected from the medical records: age; sex; pre-morbid modified Rankin scale (mRS) scores; comorbidities including hypertension, diabetes mellitus, and dyslipidemia based on the diagnoses before EVT or medications for these conditions; history of subarachnoid hemorrhage (SAH); periprocedural antiplatelet drug (aspirin, cilostazol, clopidogrel, prasugrel and their combinations) administration; aneurysm morphologies (size which is defined as the largest diameter of the aneurysm, location, type [saccular or dissecting], and status [ruptured or unruptured]); World Federation of Neurological Surgeons (WFNS) grade in patients with ruptured aneurysms ^12^; and, external ventricular drainage (EVD) installation before EVT.

### Endovascular treatment for intracranial aneurysms and intraprocedural rupture

EVT was performed under general anesthesia and systemic heparinization at any time between sheath insertion and first coil placement using techniques including simple coil embolization, double-catheter technique, and balloon- or stent-assisted coil embolization. Parent artery occlusion (PAO) was performed in the patients with ruptured dissecting aneurysms. IPR was defined as a microguidewire, microcatheter, or coil located beyond the confines of the aneurysm sac with or without angiographic contrast material extravasation, including angiographic extravasation triggered by contrast material injection during EVT ^6, 9^. When IPR was observed, rapid occlusion of the aneurysm was performed using coils, if possible. Data on the management of IPR, including heparin reversal using protamine sulfate and administration of antihypertensive medication for medical management, opening of EVD to maintain intracranial pressure, and hemostasis by inflation of a balloon microcatheter navigated around the aneurysm neck or balloon-guiding catheter (BGC) for endovascular management, were also collected. The causes of perforation, such as the framing coil, filling/finishing coil, microcatheter, microguidewire, injection of contrast agents, and hemodynamic response during IPR (defined as a 10% elevation of systolic blood pressure or heart rate from baseline) were also collected.

### Outcomes

The postoperative angiographic occlusion status was assessed using the Raymond–Roy classification: Class I, complete obliteration; Class II, residual neck; and Class III, residual aneurysm ^13^. The imaging outcomes were evaluated for hemorrhagic and ischemic lesions on computed tomography (CT) and magnetic resonance imaging (MRI), respectively. We assessed the hemorrhagic state in the subarachnoid, parenchymal, and intraventricular spaces in three stages: no change, increase, and significant increase, based on CT immediately after EVT compared with preoperative CT. We defined hemorrhage by a significant increase in any of these spaces as “increased hemorrhage.” MRI was performed to assess postoperative ischemic lesions, and the size and number of hyperintense lesions (HIL) were measured by diffusion-weighted imaging (DWI). We defined “DWI positive” as more than 10 HILs sized <5 mm, or one or more HILs sized ≥10 mm, with reference to previous studies ^14–16^. Data on aneurysm rebleeding within 2 weeks of EVT were also collected. Neurological outcomes were evaluated at discharge using mRS scores. Patients who underwent EVT for ruptured aneurysms were dichotomized into groups as good (mRS ≤3) or poor (mRS ≥4) ^17^. In patients with unruptured aneurysms, good outcomes were defined as mRS =0 or the same score as the premorbid mRS, and poor outcomes were defined as mRS worsening by 1 or more.

We first analyzed the baseline clinical characteristics of IPR. Second, we assessed the factors associated with poor outcomes in patients who underwent IPR during EVT for ruptured aneurysms and investigated the impact of hemorrhage and ischemic complications due to IPR on patient outcomes. Third, the safety and efficacy of IPR management were evaluated, focusing on controlling hemorrhage and ischemic complications.

### Statistical Analysis

We defined the following four analysis sets: (i) patients who underwent IPR; (ii) patients who underwent IPR with ruptured aneurysms; (iii) patients who underwent IPR with unruptured aneurysms; and (iv) patients who underwent IPR with ruptured aneurysms and a additionally underwent postoperative MRI.

Baseline characteristics are represented as median and interquartile range (IQR) for continuous variables and frequency and percentage for categorical variables. For the three analysis sets, except for the set of patients who underwent IPR, the distributions of baseline characteristics were compared between good/poor outcomes using Student t-test for continuous variables and chi-square test for categorical variables.

First, we assessed the effects of these factors on the good/poor outcomes in patients who underwent IPR for ruptured aneurysms. We estimated odds ratios (OR), 95% confidence intervals (CI), and p-values using univariate and multivariate logistic regression models that included age, sex, premorbid mRS, WFNS grade, and increased hemorrhage. Age was dichotomized by the median value in patients who underwent IPR with ruptured aneurysms. We also performed univariate and multivariate logistic regression analyses for good/poor outcomes in patients who underwent IPR for ruptured aneurysms and postoperative MRI. The model included age, sex, premorbid mRS score, WFNS grade, increased hemorrhage, and DWI positivity. Second, we evaluated the effects of these factors on increased hemorrhage in patients who underwent IPR for ruptured aneurysms. Univariate and multivariate logistic regression analyses were performed for age, sex, heparin reversal, antihypertensive medication, EVD opening, BGC balloon inflation, and neck balloon inflation. Finally, we investigated the effect of these factors on DWI positivity in patients who underwent IPR for ruptured aneurysms and postoperative MRI. Univariate and multivariate logistic regression analyses were also conducted using age, sex, heparin reversal, antihypertensive medication, EVD opening, BGC balloon inflation, and neck balloon inflation as covariates. Statistical significance was set at p < 0.05. All the analyses were performed using SAS version 9.4 (SAS Institute Inc., Cary, NC, USA).

## Results

### Patient Characteristics

Based on the EVT registry, we identified 3269 endovascular procedures for intracranial aneurysms (1633 endovascular procedures for unruptured aneurysms and 1636 endovascular procedures for ruptured aneurysms); IPR was observed in 74 patients, comprising 19 patients (1.16%) with unruptured aneurysms and 55 patients (3.36%) with ruptured aneurysms. The baseline clinical characteristics of the patients are shown in Table 1. The 74 patients comprised 16 (21.6%) men (median [IQR] age, 62.5 [51.0–73.0]). Sixty patients (81.1%) had premorbid mRS scores of 0. The median (IQR) aneurysm size was 4.9 mm (3.7–7.1 mm), and saccular aneurysm accounted for 79.7% (n=59). Twenty-nine patients (39.2%), including all patients with unruptured aneurysms, were pretreated with antiplatelet drugs. The most common aneurysm location was the anterior communicating artery (n=19; 25.7%), followed by the posterior communicating artery (n=16; 21.6%). The most common endovascular technique used for coil embolization was simple (n=31; 41.9%), followed by the balloon assisted one (n=29; 39.2%). Sixty-nine patients (93.2%) received intravenous heparin infusion during EVT. The most common cause of IPR was filling or finishing the coil (n=26, 35.1%), followed by framing the coil (n=23, 31.1%). The most common management for IPR was heparin reversal in 50 patients (67.6%), followed by antihypertensive medication in 42 patients (56.8%), opening of the EVD in 28 patients (37.8%) among the 30 patients who underwent EVD installation before EVT for ruptured aneurysms, inflation of the BGC in 9 patients (12.2%), and inflation of the neck balloon microcatheter in 41 patients (55.4%). PAO was performed in 12 patients, including 4 who underwent unplanned PAO for bleeding control of IPR. Twenty-six patients (35.1%) showed increased hemorrhage on CT immediately after EVT, including one patient who could not undergo postoperative CT because the patient was in a state of clinical brain death due to massive and uncontrollable IPR. Sixty-six patients underwent postoperative MRI, excluding 8 patients who were in a state of clinically close to brain death, and 32 patients (48.5%) were DWI-positive. All the patients underwent MRI within 7 days of EVT and 29 (90.6%) underwent MRI within 2 days of EVT. Thirty-six (48.6 %) patients had poor outcomes. Rebleeding within 2 weeks of EVT was observed in 7 patients (9.5%), and their occlusion status was as follows: Raymond–Roy Class I, 2; Class II, 2; Class III, 3; PAO,1.

**Table 1.** Patient characteristics of those who experienced IPR during EVT for intracranial.

|  | Overall (N=74) |
| --- | --- |
| Age | 62.5 (51.0, 73.0) |
| Male sex | 16 (21.6%) |
| Pre-morbid mRS =0 | 60 (81.1%) |
| Comorbidities and pre-medication |  |
| Hypertension | 36 (48.6%) |
| Diabetes mellitus | 5 (6.8%) |
| Dyslipidemia | 14 (18.9%) |
| SAH | 8 (10.8%) |
| Antiplatelet drugs | 29 (39.2%) |
| Aneurysm morphology |  |
| Size (mm) | 4.9 (3.7, 7.1) |
| Location |  |
| Anterior communicating artery | 19 (25.7%) |
| Posterior communicating artery | 16 (21.6%) |
| Basilar artery | 10 (13.5%) |
| Vertebral artery | 8 (10.8%) |
| Other anterior circulation aneurysm | 14 (18.9%) |
| Other posterior circulation aneurysm | 7 (9.5%) |
| Saccular aneurysm | 59 (79.7%) |
| Status of ruptured aneurysm | 55 (74.3%) |
| Endovascular technique |  |
| Simple coil embolization | 31 (41.9%) |
| Double-catheter technique | 7 (9.5%) |
| Balloon-assisted coil embolization | 29 (39.2%) |
| Stent-assisted coil embolization | 4 (5.4%) |
| Others | 3 (4.1%) |
| Heparin use | 69 (93.2%) |
| Cause of perforation |  |
| Framing coil | 23 (31.1%) |
| Filling/finishing coil | 26 (35.1%) |
| Microcatheter | 12 (16.2%) |
| Microguidewire | 4 (5.4%) |
| Injection of contrast agents | 6 (8.1%) |
| Others | 3 (4.1%) |
| Management for intraoperative rupture |  |
| Heparin reversal | 50 (67.6%) |
| Antihypertensive medication | 42 (56.8%) |
| Opening of EVD | 28 (37.8%) |
| BGC balloon inflation | 9 (12.2%) |
| Neck balloon inflation | 41 (55.4%) |
| Hemodynamic response | 37 (50.0%) |
| Angiographic occlusion status |  |
| Raymond–Roy Class 1 | 34 (45.9%) |
| Raymond–Roy Class 2 | 21 (28.4%) |
| Raymond–Roy Class 3 | 7 (9.5%) |
| Parent artery occlusion | 12 (16.2%) |
| Imaging outcomes |  |
| Increased hemorrhage | 26 (35.1%) |
| DWI positive* | 32 (48.5%) |
| Rebleeding within 2 weeks from EVT | 7 (9.5%) |
| mRS at discharge |  |
| 0 | 16 (21.6%) |
| 1 | 12 (16.2%) |
| 2 | 6 (8.1%) |
| 3 | 4 (5.4%) |
| 4 | 8 (10.8%) |
| 5 | 15 (20.3%) |
| 6 | 13 (17.6%) |

\*; Data are available for 66 participants who underwent MRI after EVT.
Abbreviations: BGC, balloon-guiding catheter; DWI, diffusion-weighted imaging; EVD, external ventricular drainage; mRS, modified Rankin Scale; SAH, subarachnoid hemorrhage

### Factors Associated with Poor Outcomes Following IPR in Patients Who Underwent EVT for Ruptured Aneurysms

The baseline clinical characteristics of 55 patients who underwent EVT for ruptured aneurysms, dichotomized according to patient outcomes, are shown in Table 2. Of these, 23 patients (41.8%) had good outcomes (mRS 0–3) and 32 patients (58.2%) had poor outcomes (mRS 4-6). Poor outcomes were significantly associated with older age, a lower percentage of pre-morbid mRS =0, a higher percentage of WFNS grades IV and V, and a higher percentage of pre-embolization EVD installation. Regarding the management of IPR, patients with poor outcomes showed a significantly higher percentage of antihypertensive medication use (65.6% vs. 34.8%, p=0.031) and opening of the EVD (68.8% vs. 26.1%, p=0.003). Regarding imaging outcomes, there was no statistically significant difference in increased hemorrhage between patients (40.6% vs. 17.4%, p=0.082). Postoperative MRI was performed in 49 patients, and DWI positivity was significantly higher in the poor outcome group than that in the good outcome group (73.1% vs. 34.8%, p =0.01). The baseline clinical characteristics of the 49 patients who underwent EVT for ruptured aneurysms and postoperative MRI, dichotomized into patient outcomes, are shown in Supplementary Table 1. The baseline clinical characteristics of the 19 patients who underwent EVT for unruptured aneurysms, dichotomized according to patient outcomes, are shown in Supplementary Table 2. All patients with unruptured aneurysms were premedicated with antiplatelet medications, and heparin infusion was administered during EVT. There were no statistically significant differences in the management of IPR and imaging outcomes between patient outcomes.

**Table 2.**
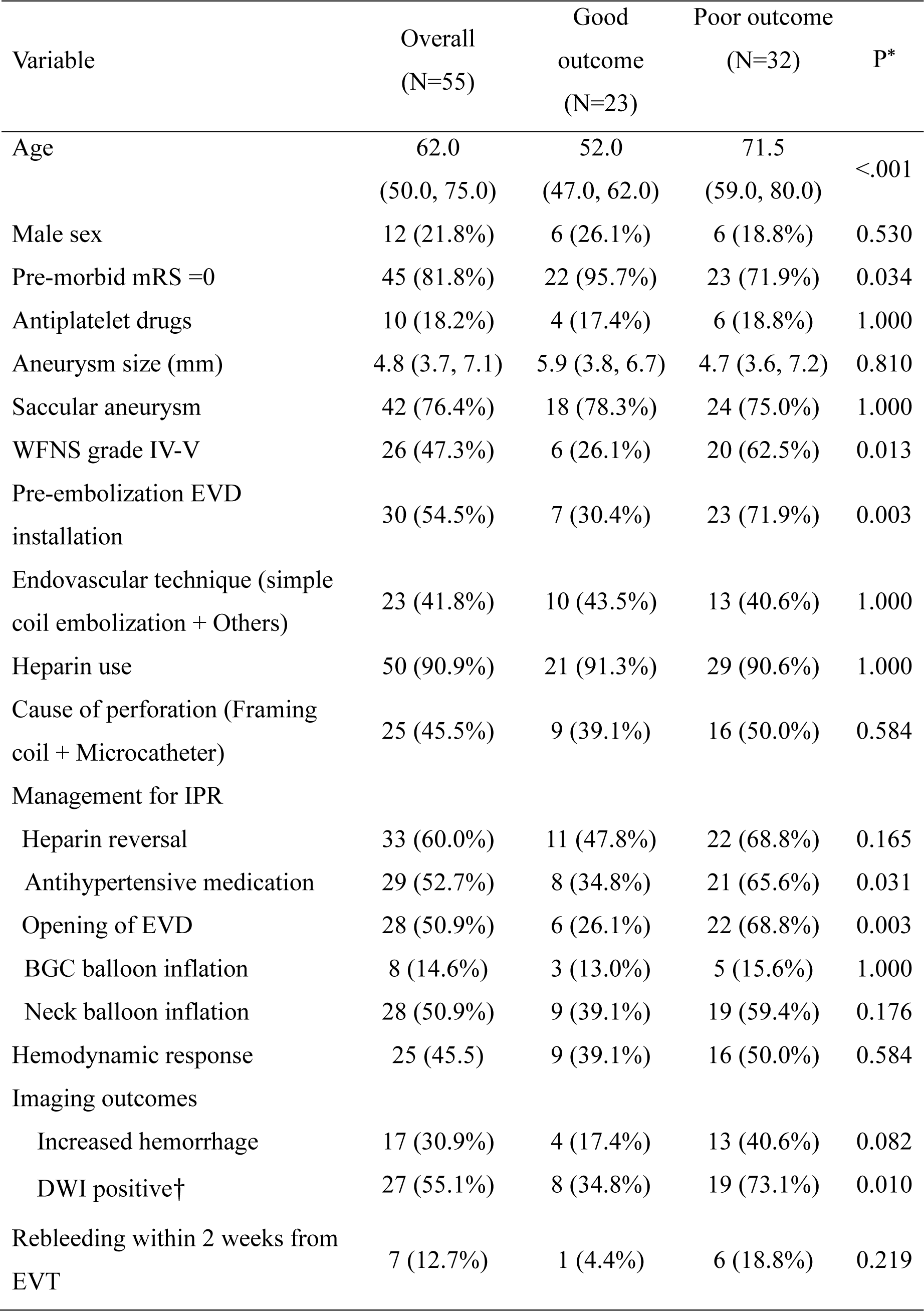
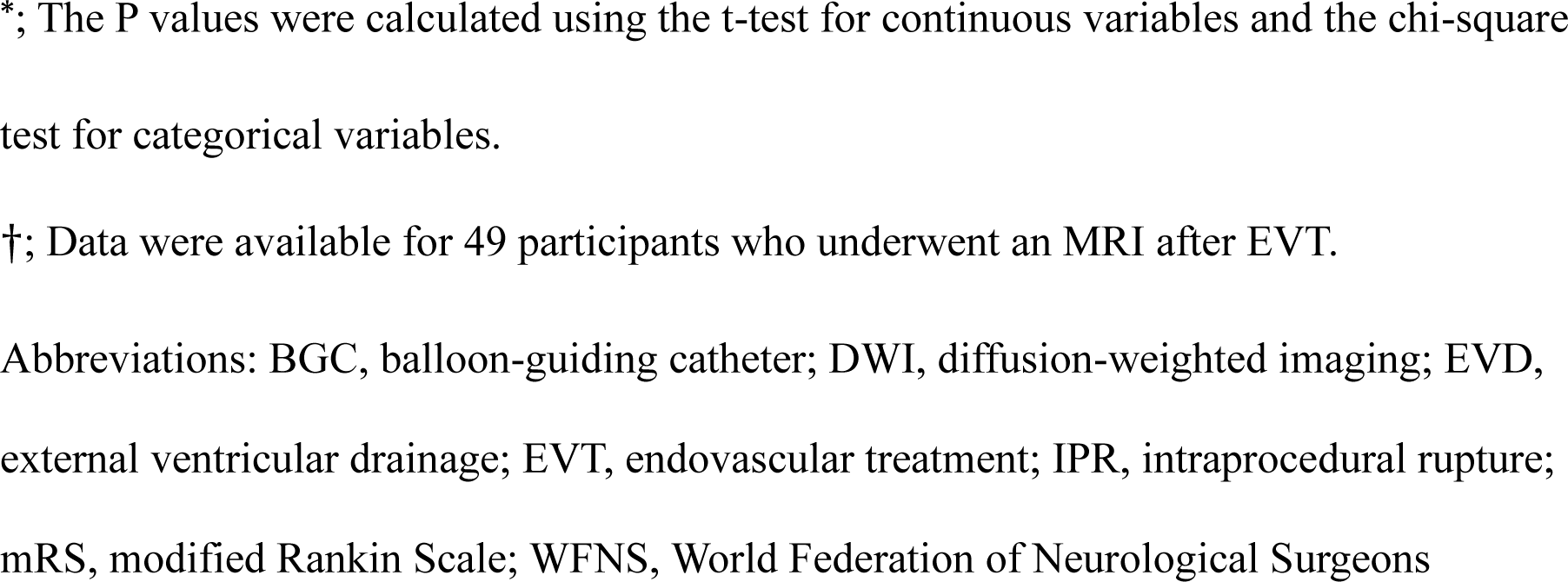
The baseline clinical characteristics of the 55 patients who underwent EVT for ruptured aneurysm dichotomized by patient outcomes.

| Variable | Overall<br>(N=55) | Good<br>outcome<br>(N=23) | Poor outcome<br>(N=32) | P* |
| --- | --- | --- | --- | --- |
| Age | 62.0<br>(50.0, 75.0) | 52.0<br>(47.0, 62.0) | 71.5<br>(59.0, 80.0) | <.001 |
| Male sex | 12 (21.8%) | 6 (26.1%) | 6 (18.8%) | 0.530 |
| Pre-morbid mRS =0 | 45 (81.8%) | 22 (95.7%) | 23 (71.9%) | 0.034 |
| Antiplatelet drugs | 10 (18.2%) | 4 (17.4%) | 6 (18.8%) | 1.000 |
| Aneurysm size (mm) | 4.8 (3.7, 7.1) | 5.9 (3.8, 6.7) | 4.7 (3.6, 7.2) | 0.810 |
| Saccular aneurysm | 42 (76.4%) | 18 (78.3%) | 24 (75.0%) | 1.000 |
| WFNS grade IV-V | 26 (47.3%) | 6 (26.1%) | 20 (62.5%) | 0.013 |
| Pre-embolization EVD<br>installation | 30 (54.5%) | 7 (30.4%) | 23 (71.9%) | 0.003 |
| Endovascular technique (simple<br>coil embolization + Others) | 23 (41.8%) | 10 (43.5%) | 13 (40.6%) | 1.000 |
| Heparin use | 50 (90.9%) | 21 (91.3%) | 29 (90.6%) | 1.000 |
| Cause of perforation (Framing<br>coil + Microcatheter) | 25 (45.5%) | 9 (39.1%) | 16 (50.0%) | 0.584 |
| Management for IPR |  |  |  |  |
| Heparin reversal | 33 (60.0%) | 11 (47.8%) | 22 (68.8%) | 0.165 |
| Antihypertensive medication | 29 (52.7%) | 8 (34.8%) | 21 (65.6%) | 0.031 |
| Opening of EVD | 28 (50.9%) | 6 (26.1%) | 22 (68.8%) | 0.003 |
| BGC balloon inflation | 8 (14.6%) | 3 (13.0%) | 5 (15.6%) | 1.000 |
| Neck balloon inflation | 28 (50.9%) | 9 (39.1%) | 19 (59.4%) | 0.176 |
| Hemodynamic response | 25 (45.5) | 9 (39.1%) | 16 (50.0%) | 0.584 |
| Imaging outcomes |  |  |  |  |
| Increased hemorrhage | 17 (30.9%) | 4 (17.4%) | 13 (40.6%) | 0.082 |
| DWI positive† | 27 (55.1%) | 8 (34.8%) | 19 (73.1%) | 0.010 |
| Rebleeding within 2 weeks from<br>EVT | 7 (12.7%) | 1 (4.4%) | 6 (18.8%) | 0.219 |

\*; The P values were calculated using the t-test for continuous variables and the chi-square test for categorical variables.
†; Data were available for 49 participants who underwent an MRI after EVT.
Abbreviations: BGC, balloon-guiding catheter; DWI, diffusion-weighted imaging; EVD, external ventricular drainage; EVT, endovascular treatment; IPR, intraprocedural rupture; mRS, modified Rankin Scale; WFNS, World Federation of Neurological Surgeons

Table 3 shows the results of the univariate and multivariate logistic regression analyses of poor outcomes among patients who underwent EVT for ruptured aneurysms. In the univariate analyses, age ≥62 years, pre-morbid mRS >0, and WFNS grades IV and V were significantly associated with poor outcomes (OR, 5.84 [95% CI, 1.80–18.93], p=0.003 for age ≥62; OR, 8.61 [95% CI, 1.01–73.69], p=0.049 for pre-morbid mRS >0; OR, 4.72 [95% CI, 1.45–15.28], p=0.010 for WFNS grades IV and V). In the multivariate analyses, age ≥62 years, WFNS grades IV and V, and increased hemorrhage were significantly associated with poor outcomes (OR, 8.53 [95% CI, 1.74–41.73], p=0.008 for age ≥62 years; OR, 12.73 [95% CI, 2.27–71.34], p=0.004 for WFNS grades IV and V; OR, 6.67 [95% CI, 1.07–41.44], p=0.042).

**Table 3.** Univariate and multivariate logistic regression analyses of poor outcomes among patients who underwent EVT for ruptured aneurysms.

| Variable | Univariate (N=55) |  | Multivariate (N=55) |  |
| --- | --- | --- | --- | --- |
|  | OR (95% CI) | P | OR (95% CI) | P |
| Age ≥62 years | 5.84<br>(1.80, 18.93) | 0.003 | 8.53<br>(1.74,41.73) | 0.008 |
| Male sex | 0.65<br>(0.18, 2.37) | 0.517 | 1.01<br>(0.20, 5.01) | 0.989 |
| Pre-morbid mRS>0 | 8.61<br>(1.01, 73.69) | 0.049 | 11.94<br>(0.85, 167.78) | 0.066 |
| WFNS grade IV and V | 4.72<br>(1.45, 15.28) | 0.010 | 12.73<br>(2.27, 71.34) | 0.004 |
| Increased hemorrhage | 3.25<br>(0.90, 11.79) | 0.073 | 6.67<br>(1.07, 41.44) | 0.042 |

Abbreviations: CI, confidence interval; DWI, diffusion-weighted imaging; mRS, modified
Rankin Scale; OR, odds ratio; WFNS, World Federation of Neurological Surgeons

The results of univariate and multivariate logistic regression analyses of poor outcomes among patients who underwent EVT for ruptured aneurysms and postoperative MRI are shown in Table 4. In the univariate analyses, age ≥62 years, WFNS grades IV and V, and DWI positivity were significantly associated with poor outcomes (OR, 6.20 [95% CI, 1.79– 21.46], p=0.004 for age ≥62 years; OR, 4.53 [95% CI, 1.34–15.37], p=0.015 for WFNS grades IV and V; OR, 5.09 [95% CI, 1.50–17.23], p=0.009 for DWI positivity). In the multivariate analyses, age ≥62 years, pre-morbid mRS >0, and WFNS grades IV and V were significantly associated with poor outcomes (OR, 8.97 [95% CI, 1.41–57.15], p=0.020 for age ≥62; OR, 19.18 [95% CI, 1.11–332.81], p=0.042 for pre-morbid mRS >0; OR, 10.78 [1.61–72.31], p=0.014 for WFNS grades IV and V).

**Table 4.** Univariate and multivariate logistic regression analyses showed poor outcomes among patients who underwent EVT for ruptured aneurysms and postoperative MRI.

| Variable | Univariate (N=49) |  | Multivariate (N=49) |  |
| --- | --- | --- | --- | --- |
|  | OR (95% CI) | P | OR (95% CI) | P |
| Age ≥62 years | 6.20<br>(1.79, 21.46) | 0.004 | 8.97<br>(1.41, 57.15) | 0.020 |
| Male sex | 0.85<br>(0.23, 3.13) | 0.807 | 1.47<br>(0.26, 8.37) | 0.664 |
| Pre-morbid mRS>0 | 6.60<br>(0.73, 59.62) | 0.093 | 19.18<br>(1.11, 332.81) | 0.042 |
| WFNS grade IV and V | 4.53<br>(1.34, 15.37) | 0.015 | 10.78<br>(1.61, 72.31) | 0.014 |
| Increased hemorrhage | 2.51<br>(0.65, 9.67) | 0.180 | 3.74<br>(0.52, 26.85) | 0.190 |
| DWI positive | 5.09<br>(1.50, 17.23) | 0.009 | 2.87<br>(0.54, 15.18) | 0.215 |

Abbreviations: CI, confidence interval; DWI, diffusion-weighted imaging; mRS, modified Rankin Scale; OR, odds ratio; WFNS, World Federation of Neurological Surgeons

### Safety and Efficacy of Management Techniques of IPR

Table 5 shows the results of the univariate and multivariate logistic regression analyses of increased hemorrhage for each variable. In the univariate analyses, antihypertensive medication use was significantly associated with increased hemorrhage (OR, 12.86 [95% CI, 2.55–64.70], p=0.002). Similar results were also observed in the multivariate analysis (OR, 13.17 [95% CI, 2.26–76.69], p=0.004 for antihypertensive medication).

**Table 5.** Univariate and multivariate logistic regression analyses of increased hemorrhage.

| Variable | Univariate (N=55) |  | Multivariate (N=55) |  |
| --- | --- | --- | --- | --- |
|  | OR (95% CI) | P | OR (95% CI) | P |
| Age≥62 years | 1.83<br>(0.56, 5.97) | 0.314 | 1.00<br>(0.20, 5.16) | 0.997 |
| Male sex | 0.37<br>(0.07, 1.93) | 0.240 | 0.35<br>(0.04, 2.81) | 0.325 |
| Management for IPR |  |  |  |  |
| Heparin reversal | 0.66<br>(0.21, 2.09) | 0.476 | 0.43<br>(0.09, 1.94) | 0.273 |
| Antihypertensive medication | 12.86<br>(2.55, 64.70) | 0.002 | 13.17<br>(2.26, 76.69) | 0.004 |
| Opening of EVD | 1.12<br>(0.36, 3.54) | 0.840 | 0.97<br>(0.23, 4.09) | 0.965 |
| BGC balloon inflation | 1.41<br>(0.30, 6.74) | 0.664 | 0.71<br>(0.10, 5.02) | 0.735 |
| Neck balloon inflation | 1.59<br>(0.50, 5.05) | 0.434 | 1.33<br>(0.32, 5.60) | 0.694 |

Abbreviations: BGC, balloon-guiding catheter; EVD, external ventricular drainage; IPR, intraprocedural rupture

The results of the univariate and multivariate logistic regression analyses with DWI positivity for each variable among patients who underwent EVT for ruptured aneurysms and postoperative MRI are shown in Table 6. In the univariate analyses, age ≥62 years and heparin reversal were significantly associated with DWI positivity (OR, 5.09 [95% CI, 1.50– 17.23], p=0.009 for age ≥62 years; OR, 5.06 [95% CI, 1.46–17.53], p=0.011 for heparin reversal). Similar results were also observed in the multivariate analysis (OR, 7.39 [95% CI, 1.43–38.32], p=0.017 for age ≥62; OR, 4.81 [95% CI, 1.09–21.14], p=0.038 for heparin reversal).

**Table 6.** Univariate and multivariate logistic regression analyses of DWI positivity.

|  | Univariate (N=49) |  | Multivariate (N=49) |  |
| --- | --- | --- | --- | --- |
|  | OR (95% CI) | P | OR (95% CI) | P |
| Age≥62 years | 5.09<br>(1.50, 17.23) | 0.009 | 7.39<br>(1.43, 38.32) | 0.017 |
| Male sex | 0.49<br>(0.13, 1.83) | 0.286 | 1.18<br>(0.20, 6.96) | 0.853 |
| Management for IPR |  |  |  |  |
| Heparin reversal | 5.06<br>(1.46, 17.53) | 0.011 | 4.81<br>(1.09, 21.14) | 0.038 |
| Antihypertensive medication | 0.93<br>(0.30, 2.86) | 0.897 | 0.49<br>(0.10, 2.32) | 0.369 |
| Opening of EVD | 1.56<br>(0.50, 4.85) | 0.446 | 1.17<br>(0.30, 4.62) | 0.821 |
| BGC balloon inflation | 0.56<br>(0.11, 2.83) | 0.486 | 2.63<br>(0.26, 26.54) | 0.411 |
| Neck balloon inflation | 2.10<br>(0.67, 6.60) | 0.204 | 2.19<br>(0.56, 8.58) | 0.263 |

Abbreviations: BGC, balloon-guiding catheter; EVD, external ventricular drainage; IPR, intraprocedural rupture

## Discussion

The present study provides real-world clinical data on IPR during EVT for intracranial aneurysms. Increased hemorrhage is associated with poor outcomes in patients who undergo EVT for ruptured aneurysms. In the management of IPR, heparin reversal was not associated with increased hemorrhage but was associated with the DWI positivity rate on MRI. In these cases, heparin reversal had no impact on clinical outcomes; however, the univariate analysis showed a significant difference in heparin reversal and DWI positivity, suggesting a potential association with the increase in the number of cases.

The overall IPR rate in the present study was 2.26%, which was more likely to occur during EVT of ruptured aneurysms than that of unruptured aneurysms. This is similar to the findings of previous studies on the prevalence of IPR ^4^. In patients with IPR, morbidity and mortality (mRS 4–6) occurred in 32 patients (58.2%) with ruptured aneurysms and four patients (21.1%) with unruptured aneurysms, representing the severe consequences of IPR, which are similar to those reported previously ^2^. The median size and common location were similar to those previously reported ^6, 18^. Although we suspected that complex endovascular techniques causing device friction or complicated procedures were more likely to cause IPR, adjunctive techniques were not significantly different between the two groups dichotomized by patient outcomes. Regarding the cause of perforation, coils were the most common cause of IPR, and the proportion of IPR related to the framing coil and microcatheter, which may be considered to be related to fatal bleeding, did not differ between the two groups dichotomized by patient outcome ^3^. These results suggest that IPR is not caused by a single factor but by various factors, including the endovascular techniques and devices used. In addition, rupture due to contrast agent injection alone was observed in six cases (8.1%). In 5 of the 6 cases, IPR occurred during diagnostic angiography prior to approaching the aneurysm. They were all WFNS grade V, and their clinical outcomes were mRS scores of 5 or 6, similar to that reported in a previous study ^19^. While performing EVT for severe SAH, careful procedures such as the gentle injection of contrast media are required. Postoperative aneurysm re-rupture occurred in 12% of patients who underwent EVT for ruptured aneurysms and experienced IPR, which was higher than the frequency of postoperative re-rupture among patients without IPR ^20^. This occurred regardless of the aneurysmal occlusion status. This may be attributable to the instability of the aneurysm and acceleration of hypercoagulation due to IPR, suggesting the possibility of further ischemic complications ^21, 22^. In patients with IPR in ruptured aneurysms, older age and the higher percentage of pre-morbid mRS =1or more and WFNS grades IV and V were related to poor outcomes. This might be because preoperative conditions have a significant influence on outcomes, even in the IPR population. EVD may be more common in patients with poor outcomes because EVD installation is performed before EVT tended to involve severe pre-EVT conditions.

In the present study, among patients who underwent postoperative MRI, the univariate analyses revealed that DWI positivity was significantly associated with poor outcomes. The incidence of DWI abnormalities following EVT in patients with SAH, which is also associated with patient outcomes, is reported to be 42–75% ^14, 23^. SAH may promote hypercoagulability as a systemic response; therefore, ischemic complications occur more frequently in EVT for ruptured aneurysms than that for unruptured aneurysms ^4, 24, 25^. Moreover, managing IPR through heparin reversal and hemostasis using a balloon in a hypercoagulable state may induce ischemic complications and compromise IPR control.

In the management of IPR, patients with poor outcomes showed a higher percentage of those with antihypertensive medication use. A hemodynamic response was observed in 23 of the 29 patients who used antihypertensive medication for the management of IPR. Rather than interpreting that the outcome was worse due to the use of antihypertensive medication, it is more reasonable to interpret that the patients who presented a hemodynamic response that required antihypertensive medication as a result of IPR tended to have poor outcomes. Therefore, none of the management strategies for IPR examined in this study were useful for bleeding control.

Among patients with IPR with ruptured aneurysms, older age and heparin reversal were associated with DWI positivity on postoperative MRI. Older age was significantly associated with multiple HIL owing to a higher prevalence of atherosclerotic risk factors, and the same trend was observed in the present study ^15, 26^. One of the optimal management strategies for IPR is rapid occlusion of the aneurysm ^1, 5^. Empirically, temporary hemostasis can be achieved using a balloon catheter; therefore, the use of a balloon is not meaningless ^3^. Although the presence or absence of a balloon did not affect the outcomes in this study, thromboembolic complications should be considered especially when heparin is neutralized ^5, 8^. The use of heparin during IPR had no effect on mortality and morbidity ^1^. Although protamine sulfate has a rapid onset of action to neutralize the effect of heparin, it causes hypotension due to vasodilation when administered rapidly; therefore, slow administration over 5 min is recommended ^27, 28^. If IPR can be controlled in a short time, heparin reversal may be avoided; if it seems difficult to control, heparin reversal can be considered, and heparinization can be restarted after hemostasis is achieved. Implementing these measures may suppress IPR-related ischemic complications. Optimal management varies depending on the case; therefore, implementing a uniform response should be reconsidered.

This study had several limitations. First, it had an observational retrospective design. To our knowledge, this is the largest study to investigate IPR during EVT for intracranial aneurysms. However, the sample size was small, and we did not have sufficient power to demonstrate the appropriateness of our results. Given the frequency and iatrogenic nature of IPR, prospective interventional studies are difficult to perform. Second, a uniform statistical analysis is challenging because of significant variations in patient backgrounds, such unruptured and ruptured conditions. Third, there were no data on activated clotting time before and after IPR, and the effects of heparin and its reversal could not be investigated in detail. Finally, imaging outcomes, including hemorrhage and ischemia, were evaluated using qualitative indicators. In particular, there are no reports on the DWI findings after IPR in patients with SAH. The validity of these criteria requires further investigation.

The present study showed real-world data on intraprocedural rupture during the EVT of intracranial aneurysms. Although rare, it can become a devastating complication. Conventional management of IPR may not be useful for controlling bleeding. Heparin reversal may be associated with ischemic complications rather than being useful for controlling bleeding during intraprocedural rupture. The situation of intraprocedural rupture is miscellaneous, and optimal management varies depending on the individual case; therefore, a uniform response should be avoided.

## Data Availability

The data supporting the findings of this study are available from the corresponding author upon reasonable request.

## Acknowledgments

None

## Non-standard abbreviations and acronyms

BGC: balloon-guiding catheter
CI: confidence interval
CT: computed tomography
DWI: diffusion-weighted imaging
EVT: endovascular treatment
EVD: external ventricular drainage
HIL: hyperintense lesion
IPR: intraprocedural rupture
IQR: interquartile range
MRI: magnetic resonance imaging
mRS: modified Rankin Scale
OR: odds ratio
PAO: parent artery occlusion
SAH: subarachnoid hemorrhage
STROBE: Strengthening the Reporting of Observational Studies in Epidemiology
WFNS: World Federation of Neurological Surgeons

## References

1. Cloft HJ, Kallmes DF. Cerebral aneurysm perforations complicating therapy with Guglielmi detachable coils: a meta-analysis. AJNR Am J Neuroradiol. 2002;23:1706–1709.

2. Elijovich L, Higashida RT, Lawton MT, Duckwiler G, Giannotta S, Johnston SC. Predictors and outcomes of intraprocedural rupture in patients treated for ruptured intracranial aneurysms: the CARAT study. Stroke. 2008;39:1501–1506. doi: 10.1161/strokeaha.107.504670

3. Santillan A, Gobin YP, Greenberg ED, Leng LZ, Riina HA, Stieg PE, Patsalides A. Intraprocedural aneurysmal rupture during coil embolization of brain aneurysms: role of balloon-assisted coiling. AJNR Am J Neuroradiol. 2012;33:2017–2021. doi: 10.3174/ajnr.A3061

4. Zheng Y, Liu Y, Leng B, Xu F, Tian Y. Periprocedural complications associated with endovascular treatment of intracranial aneurysms in 1764 cases. J Neurointerv Surg. 2016;8:152–157. doi: 10.1136/neurintsurg-2014-011459

5. Cho SH, Denewer M, Park W, Ahn JS, Kwun BD, Lee DH, Park JC. Intraprocedural Rupture of Unruptured Cerebral Aneurysms During Coil Embolization: A Single-Center Experience. World Neurosurg. 2017;105:177–183. doi: 10.1016/j.wneu.2017.05.147

6. Yamagami K, Hatano T, Nakahara I, Ishii A, Ando M, Chihara H, Ogura T, Suzuki K, Kondo D, Kamata T, et al. Long-term Outcomes After Intraprocedural Aneurysm Rupture During Coil Embolization of Unruptured Intracranial Aneurysms. World Neurosurg. 2020;134:e289–e297. doi: 10.1016/j.wneu.2019.10.038

7. Fan L, Lin B, Xu T, Xia N, Shao X, Tan X, Zhong M, Yang Y, Zhao B. Predicting intraprocedural rupture and thrombus formation during coiling of ruptured anterior communicating artery aneurysms. J Neurointerv Surg. 2017;9:370–375. doi: 10.1136/neurintsurg-2016-012335

8. Kunz M, Bakhshai Y, Zausinger S, Fesl G, Janssen H, Brückmann H, Tonn JC, Schichor C. Interdisciplinary treatment of unruptured intracranial aneurysms: impact of intraprocedural rupture and ischemia in 563 aneurysms. J Neurol. 2013;260:1304–1313. doi: 10.1007/s00415-012-6795-9

9. Sluzewski M, Bosch JA, van Rooij WJ, Nijssen PC, Wijnalda D. Rupture of intracranial aneurysms during treatment with Guglielmi detachable coils: incidence, outcome, and risk factors. J Neurosurg. 2001;94:238–240. doi: 10.3171/jns.2001.94.2.0238

10. Li MH, Gao BL, Fang C, Cheng YS, Li YD, Wang J, Xu GP. Prevention and management of intraprocedural rupture of intracranial aneurysm with detachable coils during embolization. Neuroradiology. 2006;48:907–915. doi: 10.1007/s00234-006-0147-3

11. Doerfler A, Wanke I, Egelhof T, Dietrich U, Asgari S, Stolke D, Forsting M. Aneurysmal rupture during embolization with Guglielmi detachable coils: causes, management, and outcome. AJNR Am J Neuroradiol. 2001;22:1825–1832.

12. Report of World Federation of Neurological Surgeons Committee on a Universal Subarachnoid Hemorrhage Grading Scale. J Neurosurg. 1988;68:985–986. doi: 10.3171/jns.1988.68.6.0985

13. Raymond J, Guilbert F, Weill A, Georganos SA, Juravsky L, Lambert A, Lamoureux J, Chagnon M, Roy D. Long-term angiographic recurrences after selective endovascular treatment of aneurysms with detachable coils. Stroke. 2003;34:1398–1403. doi: 10.1161/01.Str.0000073841.88563.E9

14. Lowe SR, Bhalla T, Tillman H, Chaudry MI, Turk AS, Turner RD, Spiotta AM. A Comparison of Diffusion-Weighted Imaging Abnormalities Following Balloon Remodeling for Aneurysm Coil Embolization in the Ruptured vs Unruptured Setting. Neurosurgery. 2018;82:516–524. doi: 10.1093/neuros/nyx240

15. Park JC, Lee DH, Kim JK, Ahn JS, Kwun BD, Kim DY, Choi CG. Microembolism after endovascular coiling of unruptured cerebral aneurysms: incidence and risk factors. J Neurosurg. 2016;124:777–783. doi: 10.3171/2015.3.Jns142835

16. Lim Fat MJ, Al-Hazzaa M, Bussière M, dos Santos MP, Lesiuk H, Lum C. Heparin dosing is associated with diffusion weighted imaging lesion load following aneurysm coiling. J Neurointerv Surg. 2013;5:366–370. doi: 10.1136/neurintsurg-2011-010225

17. Wilson DA, Nakaji P, Albuquerque FC, McDougall CG, Zabramski JM, Spetzler RF. Time course of recovery following poor-grade SAH: the incidence of delayed improvement and implications for SAH outcome study design. J Neurosurg. 2013;119:606–612. doi: 10.3171/2013.4.Jns121287

18. Stapleton CJ, Walcott BP, Butler WE, Ogilvy CS. Neurological outcomes following intraprocedural rerupture during coil embolization of ruptured intracranial aneurysms. J Neurosurg. 2015;122:128–135. doi: 10.3171/2014.9.Jns14616

19. Komiyama M, Tamura K, Nagata Y, Fu Y, Yagura H, Yasui T. Aneurysmal rupture during angiography. Neurosurgery. 1993;33:798–803. doi: 10.1227/00006123-199311000-00002

20. Sluzewski M, van Rooij WJ. Early rebleeding after coiling of ruptured cerebral aneurysms: incidence, morbidity, and risk factors. AJNR Am J Neuroradiol. 2005;26:1739–1743.

21. Lauridsen SV, Hvas CL, Sandgaard E, Gyldenholm T, Mikkelsen R, Obbekjær T, Sunde N, Tønnesen EK, Hvas AM. Thromboelastometry Shows Early Hypercoagulation in Patients with Spontaneous Subarachnoid Hemorrhage. World Neurosurg. 2019;130:e140–e149. doi: 10.1016/j.wneu.2019.06.019

22. Li K, Guo Y, Zhao Y, Xu B, Xu K, Yu J. Acute rerupture after coil embolization of ruptured intracranial saccular aneurysms: A literature review. Interv Neuroradiol. 2018;24:117–124. doi: 10.1177/1591019917747245

23. Bond KM, Brinjikji W, Murad MH, Kallmes DF, Cloft HJ, Lanzino G. Diffusion-Weighted Imaging-Detected Ischemic Lesions following Endovascular Treatment of Cerebral Aneurysms: A Systematic Review and Meta-Analysis. AJNR Am J Neuroradiol. 2017;38:304–309. doi: 10.3174/ajnr.A4989

24. Fukuda H, Handa A, Koyanagi M, Lo B, Yamagata S. Association of plasma D-dimer level with thromboembolic events after endovascular coil treatment of ruptured cerebral aneurysms. J Neurosurg. 2018:1–8. doi: 10.3171/2017.7.Jns171129

25. Juvela S, Siironen J. D-dimer as an independent predictor for poor outcome after aneurysmal subarachnoid hemorrhage. Stroke. 2006;37:1451–1456. doi: 10.1161/01.Str.0000221710.55467.33

26. Kang DH, Kim BM, Kim DJ, Suh SH, Kim DI, Kim YS, Huh SK, Park J, Lee JW, Kim YB. MR-DWI-positive lesions and symptomatic ischemic complications after coiling of unruptured intracranial aneurysms. Stroke. 2013;44:789–791. doi: 10.1161/strokeaha.112.669853

27. Dhakal P, Rayamajhi S, Verma V, Gundabolu K, Bhatt VR. Reversal of Anticoagulation and Management of Bleeding in Patients on Anticoagulants. Clin Appl Thromb Hemost. 2017;23:410–415. doi: 10.1177/1076029616675970

28. Horrow JC. Protamine: a review of its toxicity. Anesth Analg. 1985;64:348–361.

